# Early Recovery Protocols Effect the Opioid Prescriptions at Discharge After Major Urologic Cancer Surgery

**DOI:** 10.1101/2020.05.24.20112243

**Authors:** Kevin M. Carnes, Ashar Ata, Theodore Cangero, Badar M. Mian

## Abstract

**Introduction:** Early recovery after surgery (ERAS) protocols are designed limit the use of opioids during in-patient stay to facilitate recovery and early discharge. However, there are conflicting reports of opioids prescribed at discharge to these patients. We wished to evaluate the effect of early recovery efforts on the opioid prescriptions given at discharge after major urologic cancer surgery.

**Methods:** We reviewed opioid prescription data from patients discharged from our facility after major urologic cancer surgery from 2016 to 2018, including cystectomy, nephrectomy (total, partial) and prostatectomy. The opioid prescriptions were normalized to hydrocodone-5 mg tablet equivalents. Multivariable analysis was performed to evaluate the effect of various factors on opioid prescriptions at discharge.

**Results:** 409 patients met the inclusion criteria, with 207 without ERAS and 202 on ERAS protocol. Potent opioid (oxycodone or hydrocodone) use was reduced from 92% to 43% and tramadol use increased from 8% to 57% (p < 0.001). Following ERAS, we noted reduction in opioid prescriptions for prostatectomy (30%, p < 0.001), cystectomy (27%, p = 0.02) and all nephrectomy procedures (32%, p < 0.001). On multivariable analysis for each procedure, ERAS protocol was a significant predictor of opioid prescriptions at discharge.

**Conclusions:** A significant decrease in the opioid prescriptions given at discharge was noted after major urologic cancer surgery with the use of ERAS protocols. There was a significant shift towards the use of less potent opioids. These findings provides a benchmark for further interventions and reduction in the outpatient opioid prescriptions after major urologic surgery.

## Introduction

An estimated 50 million surgical procedures are performed in the US annually and opioid analgesics are routinely prescribed for these patients for post-operative pain control.^1, 2^ The risk of opioid use disorder, prescription misuse, and deaths resulting from overdose has been a growing problem over the last two decades. ^3^ As of September 2018, more than 67,000 Americans had died of opioid related overdose in the prior 12 months. It is common to have excessive prescriptions of opioids after surgery, with nearly two thirds of patients reporting leftover opioids following urological surgery, which may serve as the potential source of opioid abuse. ^4^ Even a short exposure to opioids in opioid-naïve patients following minor or major surgery has been associated with de novo habitual or persistent use of opioids in 5-30% of patients. ^5, 6^

Multidisciplinary, collaborative efforts have been undertaken at many centers to develop early recovery after surgery (ERAS) protocols in conjunction with dedicated members of anesthesia team and nursing staff. The ERAS protocols limit the in-patient use of opioids in order to minimize bowel dysfunction, expedite recovery and early discharge from the hospital. While it may seem intuitive that the opioid prescriptions given at discharge should also have, this has not been the case. To the contrary, some studies have reported that patients managed under ERAS protocol may be receiving similar or higher opioid prescriptions at discharge.^7^ The association between opioid prescription at discharge and ERAS protocol has not been studied well in urologic cancer surgery. Our goal was to study whether the implementation of ERAS protocols had affected the practice of opioid prescriptions given at discharge after major urologic cancer surgeries at our center.

## Materials and Methods

We reviewed the hospital medical records of patients undergoing robotic radical prostatectomy, open or robotic radical nephrectomy, open or robotic partial nephrectomy, and open radical cystectomy at our center from 2016 to 2018. The main inclusion criteria were the availability of opioid prescriptions data at discharge, including the type of medication and number of tablets prescribed. Because electronic prescription documentation was not mandatory during the entire study period, we included only those patients who had an electronically documented opioid prescription. Patients who may have received a handwritten prescription or a mention of the prescription without details in the discharge paperwork could not be included because we could not confirm the strength or the tablet count of the prescription. Due to the similarities in the incisions, the post-operative recovery protocols and discharge instructions, patients undergoing total nephrectomy, partial nephrectomy and nephroureterectomy were included in the nephrectomy group, and subdivided into either open or robotic, depending upon the approach.

We reviewed the discharge opioid prescription pattern before and after initiating the ERAS protocol for nephrectomy, prostatectomy, and cystectomy which were implemented in early to mid-2017. A brief description of the preferred analgesic medications used in ERAS protocols for various procedures is provided in supplementary materials (Table A). The discharge instructions recommended using acetaminophen and/or ibuprofen, but no other medications were prescribed. While medications such as gabapentin or pregabalin were used for some patients as part of ERAS protocol, patient were not discharged with these medications. In order to identify any patients who may have required additional opioid prescriptions as an outpatient, we reviewed the outpatient electronic medical record system of patients on ERAS protocols for 30 days after discharge. We cataloged all the phone calls made by the patients to the office and any outpatient opioid prescriptions. Due to a change in outpatient medical record system, we could not collect similar data for patients treated before ERAS protocols. The additional outpatient opioid prescription strength and dose was at the discretion of the prescriber.

We conducted an interview survey of twelve prescribers (residents and PAs), who were responsible for discharge prescriptions, to elicit descriptive responses regarding their opioid prescribing patterns including the rationale for the amount and type of medications used is provided in the supplementary data (Table B).

Primary outcomes of interest were the number of standardized opioid tablets and the type of medications prescribed at discharge. The medications prescribed included hydrocodone with acetaminophen, oxycodone (with or without acetaminophen) and tramadol. While morphine milligram equivalents (MME) are often used to standardize the opioid prescriptions, we elected to standardize the opioid dose into standardized tablet counts because a tablet is a more clinically relevant unit for the prescribers. MME data is provided in supplementary documents. Each prescription was converted into total MME and then into standardized tablet count of hydrocodone-5 mg equivalents. We used the standardized tablet count for data analysis and have provided MME data in supplementary data (Table C). The mean (standard deviation, ±SD) number of standardized tablets prescribed for each procedure were compared before and after ERAS.

Categorical variables were described and compared between before and after ERAS implementation periods using Chi-square tests and Fisher’s Exact tests. Continuous variables were compared via t-tests, Wilcoxon rank sum tests. Our outcome of interest was hydrocodone-5 mg equivalents each tablet and distributed on a continuous scale. Multivariable adjustment was done via linear regression and the adjusted effects, and 95% confidence intervals were reported. Analysis was performed using all available data for each surgical procedure. Statistical software STATA 15.0 was used for analysis and statistical significance was assessed based on alpha of 0.05 and 95% confidence interval of the differences.

## Results

Of the 409 patients undergoing major urologic procedures, 207 (51%) had surgery before ERAS and 202 (49%) after ERAS protocols were initiated. Robot-assisted laparoscopic approach was used in 285 cases, with 158 (55%) procedures before implementing ERAS protocol. The patient characteristics are outlined in Table 1. There was no difference in the age, gender, race and pre-operative opioid use amongst the group. There was a significant difference in the opioid medications prescribed. After ERAS, tramadol was prescribed at a significantly higher rate than hydrocodone or oxycodone (p < 0.001). These medications were standardized into hydrocodone-5 mg equivalent tablets for further analysis.

**Table 1.** Patient characteristics of 409 patients undergoing urologic cancer surgery.

|  | Pre-Eras, N (%) | Post-ERAS, N (%) | p Value |
| --- | --- | --- | --- |
| Gender |  |  | 0.13 |
| Male | 160 (77.3) | 143 (70.8 ) |  |
| Female | 47 (22.7) | 59 (29.2) |  |
| Race |  |  | 0.80 |
| White | 187 (90.3) | 186 (92.1) |  |
| Black | 12 (5.8) | 9 (4.5) |  |
| Others | 8 (3.9) | 7 (3.5) |  |
| Pre-op Narcotics |  |  | 0.10 |
| No | 195 (94.2) | 195 (96.5) |  |
| Yes | 12 (5.8) | 7 (3.5) |  |
| Opioid prescribed at discharge |  |  | < 0.001 |
| Hydrocodone | 49 (23.7) | 6 (2.9) |  |
| Oxycodone | 140 (67.6) | 93 (46.0) |  |
| Tramadol | 18 (8.7) | 103 (51.0) |  |

There was a significant decline in the mean opioid tablets prescribed at discharge for each procedure after initiating ERAS protocols (Figure 1). For open radical and partial nephrectomy procedures (> 90% with flank incision), the mean opioid tablets decreased by 38.2% (49.7 ± 15.4 to 30.7 ±18.5, p < 0.001) and for robotic nephrectomy procedures, the mean opioid tablets decreased by 34.6% (37.6 ± 14.6 to 24.6 ±1 4.3, < 0.001). For patients undergoing prostatectomy, 30% (33.0 ±9.1 to 23.1 ±8.3, p < 0.001) reduction in the mean opioid tablet count was noted. Opioid prescriptions given at discharge after cystectomy decreased by 27.2% (39.0 ± 17.1 to 28.4 ± 12.9, p= 0.02).

**Figure 1.**
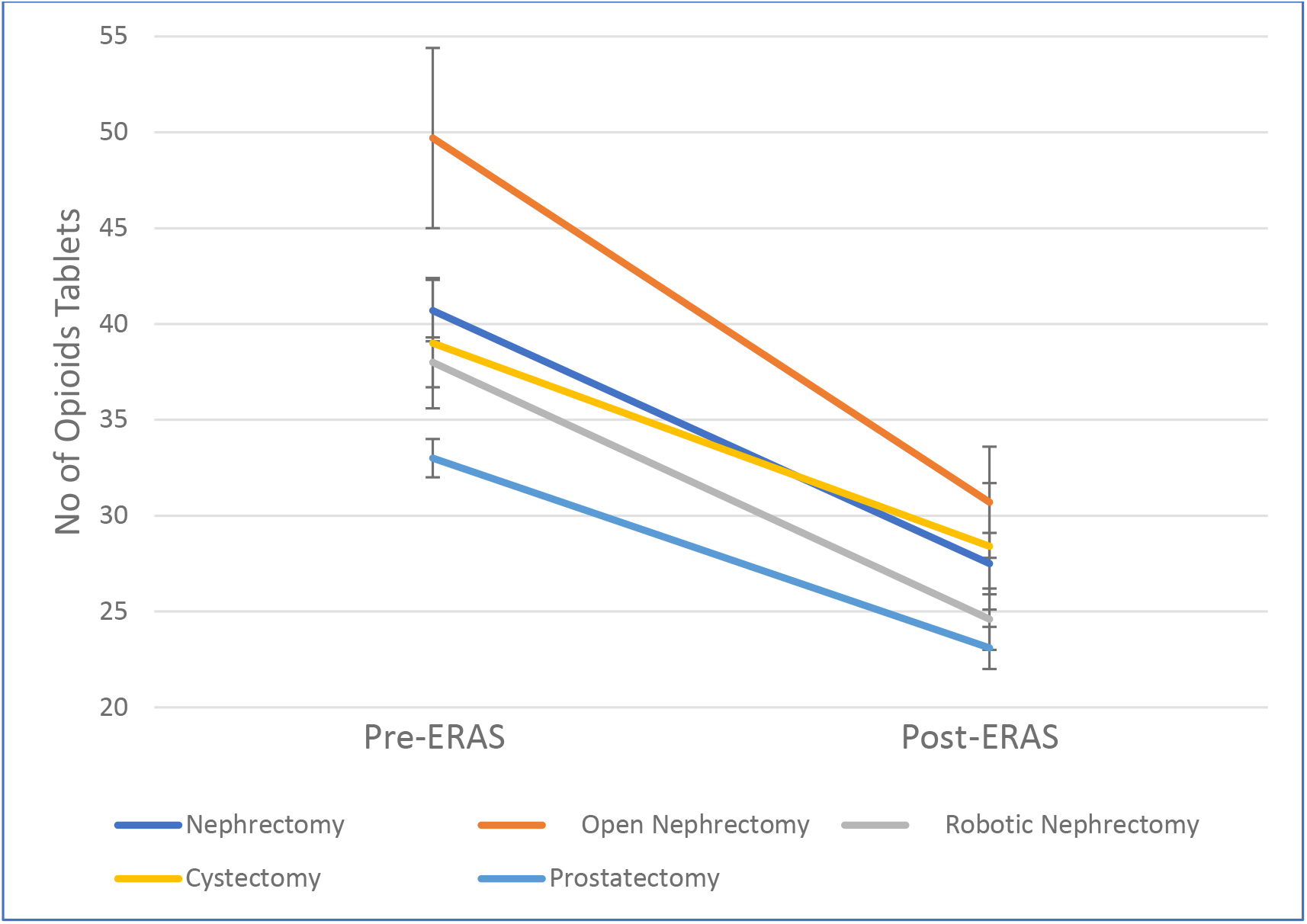
Reduction in opioid prescriptions in patients treated under the early recovery after surgery (ERAS) protocol.

We performed multivariate analysis for each of the 4 procedures individually as well including all procedures as variable. After controlling for age, gender, race and pre-operative opioid use, ERAS protocol was associated with most significant reduction in opioid prescriptions given at discharge, followed by type of surgery (Table 2). Increasing age was also associated with fewer opioid prescriptions. With open nephrectomy as the reference, other procedure types were associated with a higher rate of reduction in opioid prescriptions given at discharge. The multivariable analysis for individual procedures is provided with the supplementary data (Table D-F).

**Table 2.** Multivariable analysis of the difference in opioid prescriptions given at discharge including all procedures

|  | Difference (95%CI) | P value |
| --- | --- | --- |
| Gender |  |  |
| Female | Reference |  |
| Male | 2.75 (- 0.81, 6.32) | 0.13 |
| Age (for every 1-year increase in age) | - 0.17 (-0.30, 0.03) | 0.01 |
| Race |  |  |
| White | References |  |
| Black | -0.60 (-6.95, 5.75) | 0.85 |
| others | -1.72 (-9.25, 5.80) | 0.65 |
| Preop narcotics |  |  |
| No | Reference |  |
| Yes | 4.14 (-3.11, 11.4) | 0.26 |
| ERAS Protocol |  |  |
| No | Reference |  |
| Yes | -12.63 (-15.51, -9.75) | < 0.001 |
| Procedure type |  |  |
| Open Nephrectomy | reference |  |
| Robotic Nephrectomy | -6.85 (-10.86, -2.84) | 0.001 |
| Prostatectomy | -10.98 (-15.30, -6.67) | < 0.001 |
| Cystectomy | -4.84 (-10.49, 0.80) | 0.09 |

The opioid formulations prescribed at discharge included hydrocodone with acetaminophen, oxycodone, and tramadol (Figure 2). Before ERAS protocol, 91% of patients received either hydrocodone (24.7%) or oxycodone (67.6%). After ERAS protocol, the use of tramadol increased from 9% to 51% (p < 0.001).

**Figure 2.**
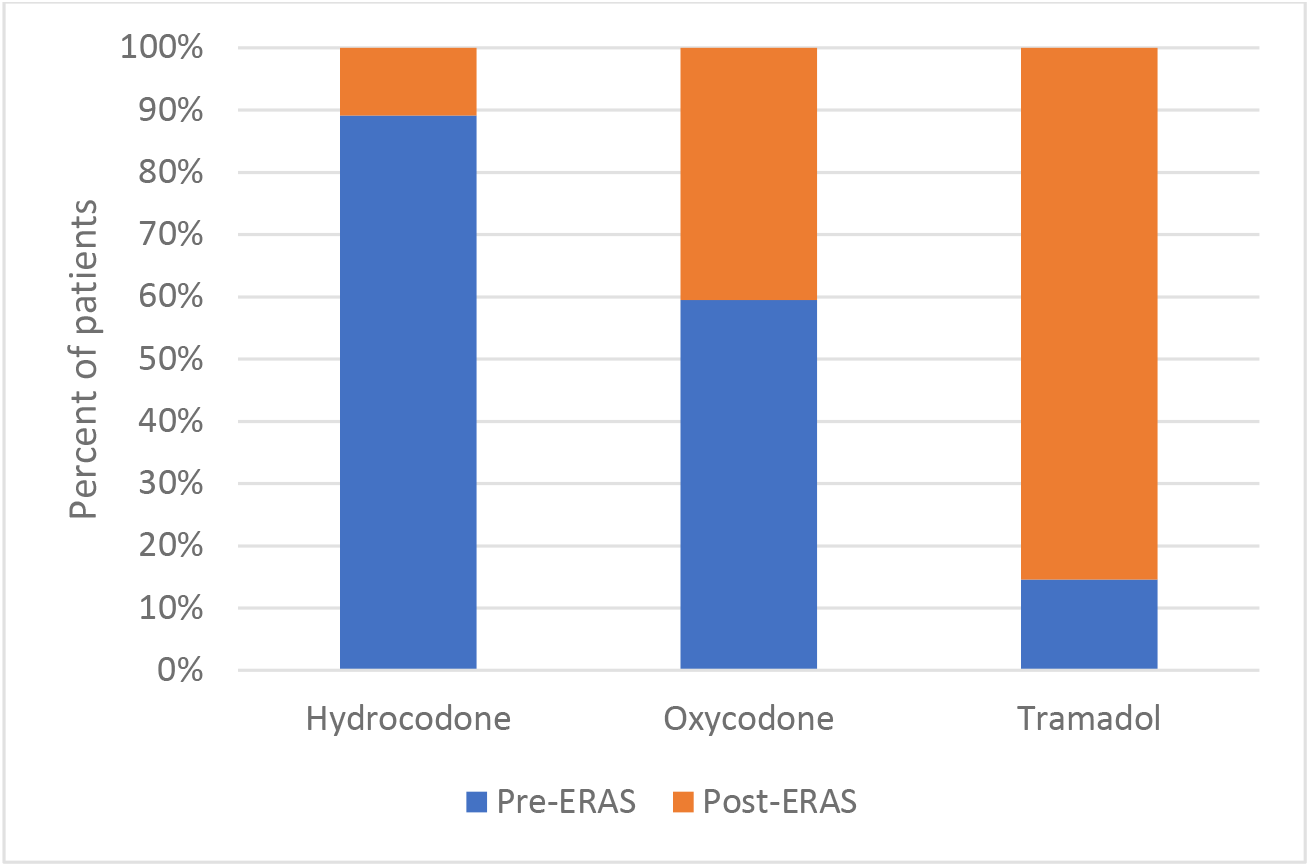
Change in the type of opioid medications prescribes at discharge from the hospital after ERAS protocol.

Upon reviewing the outpatient medical records of 202 patients on ERAS protocols, we identified phone calls from 38 (18.8%) patients within 30 days of discharge. Most of the call were related to discomfort due to Foley catheter, bladder spasms, hematuria, and GI issues. Of these, 13 (6.6%) patients were prescribed 23.0 hydrocodone-5 mg equivalent tablets (range 10-45 tablets), and 11 of 13 patients received hydrocodone or oxycodone. All 13 patients requiring additional opioids as outpatient had undergone either an open nephrectomy procedure (8 patients) or robotic nephrectomy procedure (5 patients).

## Discussion

Nearly 200 million opioid prescriptions are dispensed per year in the United States.^8^ The rate of opioid prescription is the highest amongst certain specialties including pain medicine (48.6%), surgery (36.5%), and physical medicine & rehabilitation (35.5%).^9^ In addition to illicit drugs, opioid prescriptions given by providers are a major contributor to the opioid abuse epidemic. Nearly 40% of opioid overdose-related deaths occur due to prescribed opioids.^10^ Among patients diagnosed with opiate dependence, 80% had received an opioid prescription prior to their abuse diagnosis and 51% had a family member who had an opioid prescription. ^11^ In a recent study of major procedures involving kidney or prostate in 155 patients, Theisen et al reported an average of 26 oxycodone-equivalent (or 39 hydrocodone-equivalent) tablets were prescribed per patient at discharge, 60% of which remained unused.^12^ Thus, the impact of opioids prescribed by medical professionals cannot be overstated. It’s essential to identify the trends and factors associated with opioid prescriptions given for use at home at hospital discharge. The ERAS protocols are being utilized at many centers in order to hasten recovery through reduced use of opioids during the in-patient recovery period. We wished to determine whether the opioid reduction measure used for in-patient recovery had an impact on opioid prescriptions given at discharge.

In our 409 patients undergoing abdominal surgery for urologic cancers (open or robotic), we noted a significant, 27% to 38%, decrease in opioid prescriptions for various procedures. On multivariate analysis, ERAS protocol was independently associated with decrease in outpatient opioid prescriptions. The largest decrease after initiating ERAS protocol was noted in the open nephrectomy group (from 45 to 24 hydrocodone-equivalent tablets), while a smaller, but significant, decrease was noted in the robotic-assisted prostatectomy group (from 33 to 23 hydrocodone-equivalent tablets).

It’s important to make a distinction between opioids used during inpatient stay which are dictated by the ERAS protocol and the opioid prescriptions given at discharge. The ERAS protocols are very prescriptive about use of medications during hospital stray and usually include some educational material and/or some counseling by the nurse about opioid use. However, the ERAS protocols typically did not address the opioid prescriptions given at discharge. We reviewed several different ERAS protocols for abdominal/pelvic surgery from other specialties and centers, and did not find any specific directions for opioids prescriptions to be given at discharge. It’s interesting to note that most of the measures listed under ERAS protocols, including medications, have been used by various physicians at their discretion. Formalizing the ERAS protocols, with the inclusion surgeons, anesthesiologists and nurses, brings everyone’s focus on reducing inpatients opioid use for early discharge. It necessitates daily discussion and education of the patients and prescribers about opioids, bringing a cultural shift within the medical team. It’s like that this enhanced focus on reducing opioids during recovery resulted in reduction of opioid prescriptions given at discharge for our patients

There are conflicting reports in the literature about opioid prescriptions at discharge for patients managed with ERAS protocols. Brandal et al reviewed colorectal surgery procedures and reported that there was no decrease in the opioid prescriptions at discharge when compared to the pre-ERAS cohort. Unexpectedly, even the subgroup with low pain scores during hospitalization and no perioperative opioid use were discharged with similarly high rate of opioid prescriptions.^13^ Another study of the opioid prescribing patterns after colorectal surgery in 2018 demonstrated that while ERAS protocol resulted in a shorter hospital stay, these patients were prescribed significantly more opioids at discharge than the patients not on the ERAS protocol (61.4 vs. 48.4 hydrocodone-5 mg equivalents).^7^ These surprising findings suggest that for many ERAS protocols, the emphasis remains on early discharge from the hospital and not necessarily on reducing opioid dissemination. In a report from 2019, Patel et al implemented standardized post-operative prescribing guidelines (separate from the inpatient recovery guidelines) which resulted in reduced post-prostatectomy opioid prescriptions from 224 MME to 120 MME (from 45 to 24 hydrocodone-5 mg equivalents) which is similar to opioids prescribed to our prostatectomy patients on ERAS (from 33 to 23 tablets) at discharge.^14^ Interestingly, despite minimally invasive prostatectomy procedure, their pre-intervention patients were receiving relatively large amount of opioid prescriptions. In these studies referenced above, the invasiveness of the procedure and early discharge did not correlate with opioid prescribing patterns. In our cohort, we demonstrated a significant decrease in opioid prescriptions after discharge for both the open, radical as well as minimally invasive cancer surgery of kidney, bladder and prostate.

One of the concerns with limiting opioid prescriptions at discharge is that it could result in poor pain control, excessive number of phone calls to the office and need for additional opioid prescriptions. While nearly 18% of patients made a phone call to office, just over 6% needed additional opioid prescriptions, primarily in open nephrectomy patients. Further, 9 of the 13 patients requiring additional opioids were age 55 or younger. While we do not have comparative data, the age group and opioid requirements are in line with the publishes literature. These findings identify a subgroup of patients that could benefit from adjustments in post-operative instructions and pain medications such as opioid doses and additional medications such as gabapentin or topical analgesics.

Our interview survey of the prescribers revealed several informative aspects of their prescribing patterns shown in supplementary data (Table B). All prescribers were aware of the opioid abuse crisis and affirmed that they had been prescribing very few and less-potent opioids at discharge. Our ERAS protocols listed tramadol as the preferred oral opioid to be used during hospitalization. Most prescribers viewed tramadol to be associated with a lower potential risk of abuse. This view likely resulted in over 5-fold increase in tramadol and 88% reduction in hydrocodone prescriptions given at discharge for all procedures. When asked about using tablet counts and morphine milligram equivalents (MME) to decide on the number and strength of the prescription, none of the twelve prescribers have ever used MME in practice. We believe that the opioids data should be presented in tablet equivalents because it is clinically more familiar and translatable unit for the readers (MME data is provided in supplementary table C).

Unused opioids that are usually kept by the patients after surgery pose a significant risk of opiate misuse. Cabo et al found that 79% of patients who received an opiate prescription after urological surgery had leftover medication. ^15^ In another study, pharmaceutical opioid diversion was associated with 63% of opioid-related deaths, with diversion prevalence being the highest among the 18 -24 year old decedents. ^16^ In the studies by Patel and Thiessen, robotic prostatectomy patients were given 24 and 39 hydrocodone-5 mg equivalent tablets, of which 68% and 60% remained unused, respectively.^12, 14^ Our robotic prostatectomy patients received 23 hydrocodone-5 mg equivalent tablets. While we do not have that specific data for our cohort, it’s likely that the rate of unused tablets would be similar.

To our knowledge, this is one of the only studies of urologic cancer surgery demonstrating a reduction in opioid prescriptions at discharge for patients managed with ERAS protocol after our most frequent major surgery (both open and robotic). Our study has other limitations which are associated with a retrospective analysis. We included only those cases where complete information about the opioid dose and strength was documented. It’s possible that the excluded patients could have received opioid prescription at discharge or after discharge through another physician or outpatient clinic. The reduction in opioid prescriptions given at discharge for ERAS patients did not results in an excessive need for additional outpatient prescriptions. However, the data for the non-ERAS group was unavailable and incomplete so that a meaningful comparison between the groups was not possible. While we demonstrated a reduction in opioid prescriptions written at discharge, published reports suggest that further reduction may be possible since up to 60 % of prescribed opioids may remain unused.^12,14^ Further efforts, such as adjusting the ERAS protocol and educational materials, made in reducing the dissemination of excessive prescription opioids and abuse potential are of utmost importance in curbing the opioid abuse epidemic.

## Conclusions

We noted a significant decrease in opioid prescriptions given at discharge after major, open or robotic, urologic surgery for patients managed with ERAS protocol. There was a significant reduction not only in the total opioid dose but also significant shift towards the use of less potent opioid medications. It’s essential to continue all efforts to mitigate the harmful effects of excessive opioid prescriptions. This data will serve as a benchmark to develop further interventions to reduce opioid prescriptions after urologic cancer surgery and limit the availability of the opioids in the community.

## Data Availability

The de-identified patient data used for this study is available

## Supplementary Data

**Supplementary Table A.** Analgesic medications used in the ERAS protocols.

|  | Cystectomy, Open | Nephrectomy, Open | Nephrectomy, Robotic | Prostatectomy, Robotic |
| --- | --- | --- | --- | --- |
| Pre-operative medications* | Acetaminophen<br>Celecoxib if GFR>60<br>Pregabalin/Gabapentin<br>Scopolamine patch | Acetaminophen<br>Celecoxib if GFR>60<br>Pregabalin/Gabapentin<br>Scopolamine patch | Acetaminophen<br>Celecoxib if GFR>60<br>Pregabalin/Gabapentin<br>Scopolamine patch | Acetaminophen<br>Celecoxib if GFR>60<br>Pregabalin/Gabapentin<br>Scopolamine patch |
| Intra-operative medications*# | Propofol; Ketamine<br>Dexmedetomidine;<br>Lidocaine; Opioids<br>Ketorolac | Propofol; Ketamine<br>Dexmedetomidine;<br>Lidocaine; Opioids | Propofol; Ketamine<br>Dexmedetomidine;<br>Lidocaine; Opioids | Propofol; Ketorolac;<br>Opioids |
| Regional analgesia | TAP Catheters | Intercostal catheters | Long acting<br>Bupivacaine | Long acting<br>Bupivacaine |
| Post-operative, inpatient | Acetaminophen;<br>Ibuprofen;<br>Tramadol; Oxycodone | Acetaminophen;<br>Ibuprofen;<br>Tramadol; Oxycodone | Acetaminophen;<br>Ibuprofen;<br>Tramadol; Oxycodone | Acetaminophen;<br>Ibuprofen;<br>Tramadol; Oxycodone |

\*Not all medications are used in each patient; At the discretion of anesthesia team; Listed in the order of preference
#All medications dosed for ideal body weight
TAP: Transversus Abdominis Plane

**Supplementary Table B.** Descriptive responses to our interview survey from prescribers who were responsible for discharge medications.

| Question | Most frequent responses* |
| --- | --- |
| Are prescription opioids a significant contributor to the opioid crisis? | Yes.<br>Significant.<br>Moderate. |
| Has your opioid prescription pattern changed in the last year?<br>Why? | Yes.<br>Prescribing fewer tablets.<br>Patients don't seem to be requiring opioids as in-patients.<br>Patients often have leftover tablets. |
| How do you decide which medication to use?<br>Why? | We have been using tramadol more frequently in the hospital.<br>It's less addictive.<br>Avoids concerns over acetaminophen dose limit since that is used first. |
| Are you familiar with morphine milligrams equivalents (MME)? | Yes.<br>Somewhat. |
| Do you sometimes use MME to standardize the dosing of your prescriptions? | Never |
| How do you decide on the amount (tablets) of opioids? | Based on in-patient pain control.<br>Out of habit.<br>Instructed by other team members.<br>Don't want the patient to run out of medications.<br>Avoid phone calls. |

\*Listed in order of frequency

**Supplementary Table C.**
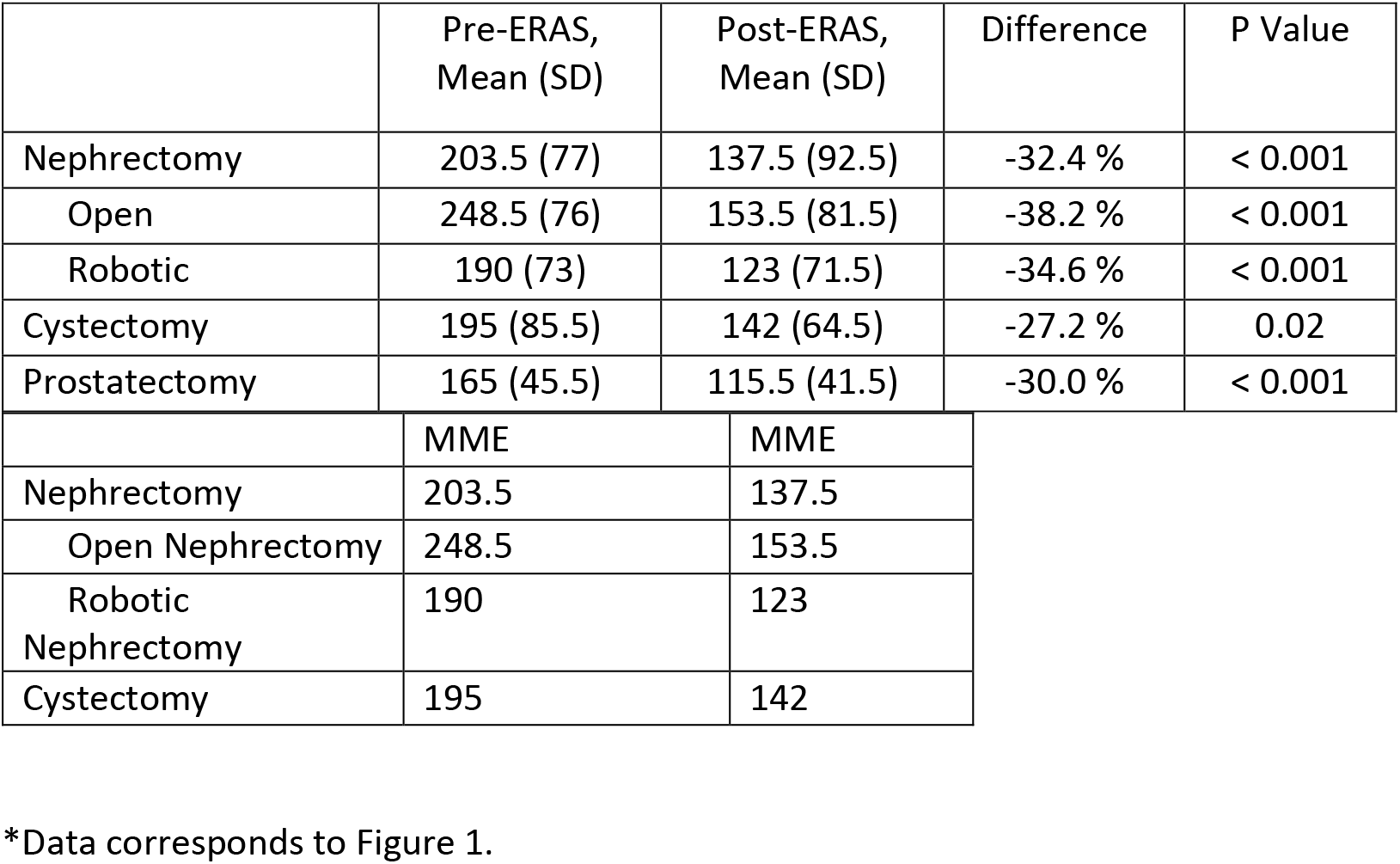
Reduction in opioid prescriptions after initiating ERAS protocols, standardized to morphine milligram equivalents^*^.

**Supplementary Table D.** Multivariable analysis of the difference in opioid prescriptions given at discharge after all nephrectomy procedures

|  | Difference (95%CI) | P value |
| --- | --- | --- |
| Gender |  |  |
| Female | Reference |  |
| Male | 3.50 (-1.06, 8.05) | 0.13 |
| Age (every 1-year increase in age) | - 0.16 (-0.34, 0.02) | 0.08 |
| Race |  |  |
| White | References |  |
| Black | 1.16 (-7.80, 10.13) | 0.79 |
| others | -0.57 (-11.01, 9.87) | .91 |
| Preop narcotics |  |  |
| No | Reference |  |
| Yes | 3.21( -7.65, 14.07) | 0.56 |
| ERAS protocol |  |  |
| Pre | Reference |  |
| Post | -14.64 (-19.29, -9.99) | < 0.001 |
| Procedure type |  |  |
| Open | reference |  |
| Robotic | -7.40 (-12.21, 2.59) | 0.003 |

**Supplementary Table E.** Multivariable analysis of the difference in opioid prescriptions given at discharge after prostatectomy

|  | Difference (95%CI) | P value |
| --- | --- | --- |
| Age (every 1-year increase in age) | -0.29 (-0.56, 0.025) | 0.03 |
| Race |  |  |
| White | References |  |
| Black | -3.72 (-11.58, 4.13) | 0.35 |
| Others | -4.49 (-13.44, 4.46) | 0.32 |
| Preop narcotics |  |  |
| No | Reference |  |
| Yes | -11.63 (-30.56, 7.28) | 0.23 |
| ERAS pathway |  |  |
| Pre-ERAS | Reference |  |
| Post-ERAS | 10.17 (-13.12, -7.22) | < 0.001 |

**Supplementary Table F.** Multivariable analysis of the difference in opioid prescriptions given at discharge after radical cystectomy

|  | Difference (95%CI) | P value |
| --- | --- | --- |
| Gender |  |  |
| Female | Reference |  |
| Male | -1.83 (-13.05, 9.38) | 0.74 |
| Age (every 1-year increase in age) | -0.71 (-0.49, 0.34) | 0.73 |
| Race |  |  |
| White | References |  |
| Black | -2.60 (-7.95, 5.40) | 0.85 |
| others | 1.12 (-9.25, 12.80) | 0.65 |
| Preop narcotics |  |  |
| No | Reference |  |
| Yes | 10.58 (-5.99, 27.17) | 0.20 |
| ERAS pathway |  |  |
| Pre | Reference |  |
| Post | -7.48 (-18.64, 3.68) | 0.18 |

## Notes

### Competing Interest Statement

The authors have declared no competing interest.

### Funding Statement

No external funding was received for this research.

### Author Declarations

This study was approved by the Institutional Review Board of Albany Medical Center.

